# Evaluation of long-term sequelae by cardiopulmonary exercise testing 12 months after hospitalization for critical COVID-19

**DOI:** 10.1101/2022.09.12.22279779

**Authors:** Sofia Noureddine, Pauline Roux-Claudé, Lucie Laurent, Ophélie Ritter, Pauline Dolla, Sinan Karaer, Frédéric Claudé, Guillaume Eberst, Virginie Westeel, Cindy Barnig

## Abstract

**Background:** Cardiopulmonary exercise testing (CPET) is an important clinical tool that provides a global assessment of the respiratory, circulatory and metabolic responses to exercise which are not adequately reflected through the measurement of individual organ system function at rest. In the context of critical COVID-19, CPET is an ideal approach for assessing long term sequalae.

**Methods:** In this prospective single-center study, we performed CPET in 60 patients, 12 months after a critical COVID-19 infection that required intensive care unit (ICU) treatment. Lung function at rest and chest computed tomography (CT) scan were also performed.

**Results:** Twelve months after severe COVID-19 pneumonia, the majority of the patients had a peak O_2_ uptake (V’O_2_) considered within normal limits. However, length of ICU stay remained an independent predictor of V’O_2_. Surprisingly, more than half of the patients with a normal peak predicted V’O_2_ showed ventilatory inefficiency during exercise (high VE/VCO_2_ ratio and high VE/VCO_2_ slope) with increased physiological dead space (VD/Vt) and low end-tidal CO_2_ partial pressure (PETCO_2_) values. This impairment was even more pronounced in patients with persistent dyspnea. Notably, peak VD/Vt values were positively correlated with peak D-Dimer plasma concentrations from blood samples collected during ICU stay.

**Conclusions:** Even if reduced exercise capacity was rare 12 months after critical COVID-19, more than half of the patients with normal exercise capacity showed ventilatory inefficiency.

## Background

In December 2019, Wuhan city identified a new type of coronavirus, named severe acute respiratory syndrome coronavirus 2 (SARS-CoV-2), that rapidly spread all over the world and caused an immense global health crisis. Most patients presented mild to moderate respiratory disease, experiencing cough, fever, headache, myalgia, diarrhea and anosmia. However, around 3–20% of people with SARS-CoV-2 required hospitalization and a considerable subset needed intensive care because of respiratory failure with severe hypoxemia and bilateral radiographic opacities [1].

Studies found that most SARS-CoV-2 survivors, even those who were critically ill during hospital stay, have normal pulmonary function tests within 12 months after symptom onset [2]. Nevertheless, more than half of the patients who recover from Coronavirus disease 2019 (COVID-19) complain of long-term persistent dyspnea [3].

Cardiopulmonary exercise testing (CPET) is an important clinical tool that provides a global assessment of the respiratory, circulatory and metabolic responses to exercise which are not adequately reflected through the measurement of individual organ system function at rest [4]. In the context of COVID-19, CPET is an ideal approach for unmasking functional anomalies and long term sequalae. To date, only a few studies have investigated the exercise capacity in patients who have recovered from COVID-19. Assessment in the short-term post-COVID period revealed mostly a mild decrease in peak O_2_ uptake (V’O_2_) and a low anaerobic threshold, without cardiac impairment or ventilatory limitation, suggestive of physical deconditioning [5, 6]. However, there is currently limited data on long term functional capacities in patients after COVID-19, especially after critical infection.

The aim of our study was to evaluate cardiopulmonary exercise capacities in a prospective cohort of patients that required critical care management during the first wave of COVID-19, 12 months after symptom onset.

## Methods

### Study design and subjects

This was a prospective single-center observational cohort study. All patients who were admitted between April to June 2020 to any of the intensive care units (ICU) of the University Hospital of Besançon (France) for a COVID-19 infection were contacted upon hospital discharge and invited to participate in the trial. Patients were eligible if they were >18 and < 80 years old and had initially confirmed SARS-CoV-2 infection by quantitative RT-PCR on nasal swabs or bronchoalveolar lavage. Patients were excluded if they were known to have prior chronic respiratory insufficiency, if they had a significant psychiatric pathology, or if they had a life expectancy estimated at less than one year. The protocol was approved by the ethics committee (Comité de Protection des Personnes (CPP) Grand-Est Ref 21 04 11) and written informed consent was obtained from all patients at the time of enrollment.

The study consisted of a follow-up at 3, 6 and 12 months after symptom onset (NCT04519320). From 149 patients that were initially admitted to intensive care with a diagnosis of SARS-CoV-2, a total of 85 patients were included in the cohort study (suppl Figure). Seventy-three patients (86%) completed the 12 months visit. A clinical evaluation, lung function tests, chest computed tomography (CT) and CPET were carried out on the same day. A total of 64 patients performed CPET (2 patients declined to perform CPET, 2 had no negative RT-PCR control for SARS-CoV-2 and 6 had contraindications for performing CPET (pericardial effusion, acid-base disorders, orthopedic pathology and recent head trauma)). Four patients were excluded from the final analysis, 3 because of submaximal efforts and 1 patient that had presented severe arrhythmia during the test leading to an early exercise termination.

### Pulmonary function tests

Pulmonary function tests were realized in all the patients and included spirometry, measurement of lung volumes by plethysmography and single-breath determination of diffusion capacity of the lung for carbon monoxide (D_LCO_) (Platinum Elite, MGC Diagnostic Coroporation). Predicted normal values were derived from the reference values in accordance with current recommendations [7, 8]. The modified Medical Research Council (mMRC) dyspnea scale (0 to 4) was used to rate chronic dyspnea [9]. Participants were categorized as having dyspnea (mMRC ≥ 1) or not (mMRC = 0).

### CT image acquisition and analysis

Chest CT scans were acquired in the supine position at full inspiration without contrast medium (Revolution CT; GE Healthcare, Milwaukee, WI, USA). CT images were assessed by two readers blinded to clinical data that evaluated the presence and extent of ground-glass opacities (GGOs), reticulations, bronchiectasis, emphysema and honeycombing as defined by the glossary of terms of the Fleischner Society [10].

### Cardiopulmonary exercise test

Symptom-limited incremental CPET was performed according to the ERS guidelines on an electronically braked cycle ergometer (Ergometrics 900; Ergoline; Bitz, Germany) [11].

After a steady-state resting period, a 3 min warm-up was conducted at about 20% of individually estimated maximal workload. A progressive increase in workload was then applied every minute (10 to 20 W/min) depending on the patient’s physical condition, medical history and according to a total exercise time between 8 and 12 minutes. Tests were terminated at the point of symptom limitation (peak exercise) or in the presence of electrocardiographic changes. Subjects rated the magnitude of their perceived breathing and leg discomfort by pointing to a number on the 10-point Borg scale [12]. Oxygen saturation with pulse oximetry, heart rate (HR) and 12-lead electrocardiogram (ECG) and non-invasive blood pressure measurements were monitored throughout exercise.

Breath-by-breath gas exchange values were measured using a Masterscreen CPX metabolic cart (MGC-CPX System; MGC Diagnostics Corporation) and were expressed as 30 s averages, according to recommended guidelines. Minute ventilation (V’E), oxygen uptake (V’O_2_), carbon dioxide production (V’CO_2_), end-tidal partial pressure of carbon dioxide (PetCO_2_), tidal volume (Vt) and breathing rate were recorded. Oxygen pulse (V’O_2_/heart rate), ventilatory equivalent for oxygen uptake (V’E/V’O_2_) and ventilatory equivalent for carbon dioxide production (V’E/V’CO_2_) were calculated. The respiratory exchange ratio (RER) was defined as V’CO_2_/V’O_2_. The anaerobic threshold (AT) was determined by both ventilatory equivalents and V-slope methods. V’E/V’CO_2_ slope was calculated from rest to peak exercise.

Blood samples were drawn from the arterialized earlobe and measurements of partial pressure of oxygen (PaO_2_) and carbon dioxide (CO_2_) were performed at rest, at anaerobic threshold (AT) and at peak exercise. Lactatemia was determined at rest, at AT, at maximal exercise and 5 min of recovery time. Breathing reserve (BR) % was calculated as BR = (predicted maximum voluntary minute ventilation [MMV] – peak VE)/MMV x 100, with predicted MMV = FEV1 × 40. Peak heart rate (HR) was expressed as a percentage of maximum predicted HR, calculated as HR max = 210 – (0.65 × age). Physiological dead space (VD/Vt) was calculated according to Bohr’s equation corrected for the additional instrument dead space: VD/Vt = (PaCO_2_ – PetCO_2_ mean)/PaCO_2_ – (VD [machine]/ Vt).

Tests were considered maximal if a plateau of the V’O_2_ > 60 seconds was obtained (variation of V’O_2_ <150 mL between 2 increments), RER > 1.1, a perceived exertion >7 on the Borg scale, peak HR > 100% of predicted, breathing reserve <15% and/or important metabolic acidosis.

Normal predicted values for V’O_2_ were calculated according to the reference equation of Wassermann [13]. A reduced peak exercise capacity was defined by a peak V’O_2_ < 85% of predicted [4].

### Statistical analyses

Statistical analysis was performed using GraphPad Prism version 9.0 (GraphPad Software Inc., San Diego, CA, USA). Normal distribution of quantitative variables was tested by the Kolmogorov-Smirnov test. Descriptive statistics are presented as mean ± standard deviation (SD), median (25^th^ to 75^th^ percentile), or number (%), as appropriate. Student’s T or Mann–Whitney U-test tests were computed to assess statistical differences between groups for normal or non-normal quantitative variables, respectively. Categorical variables were analyzed by Fisher’s exact test when appropriate. Correlations were examined by Spearman rank test or Pearson test. Multiple linear regression was applied for peak V’O_2_ (ml/kg/min) as dependent variable, using a stepwise approach of potential determinants that showed significant associations in previous univariate analysis. Age, sex and body mass index (BMI) were included in the final multivariable model. The reported p values were two-sided, with a significance level set at p<0.05.

## Results

### Baseline characteristics of the study population

The demographics, comorbidities and ICU treatments of patients at inclusion are summarized in Table 1. The mean age was 64.6 years (± 9.6), 78% were male. All patients were initially admitted in an intensive care unit (ICU) and were treated according to local standards at that time. The majority of the patients fulfilled criteria for initial ARDS according to the Berlin definition [14], 90% were intubated, 45% received steroids and more than 90% received early anticoagulant therapy. A pulmonary embolism was diagnosed in 27% of the patients during their stay. Peak values during ICU stay for main blood laboratory findings are shown in Table 1. All patients included in the study had early rehabilitation during their hospital stay. More than half of the patients (65%) were further referred to a pulmonary rehabilitation center and most of the other patients had regular home physiotherapy sessions after their hospital discharge.

**Table 1.**
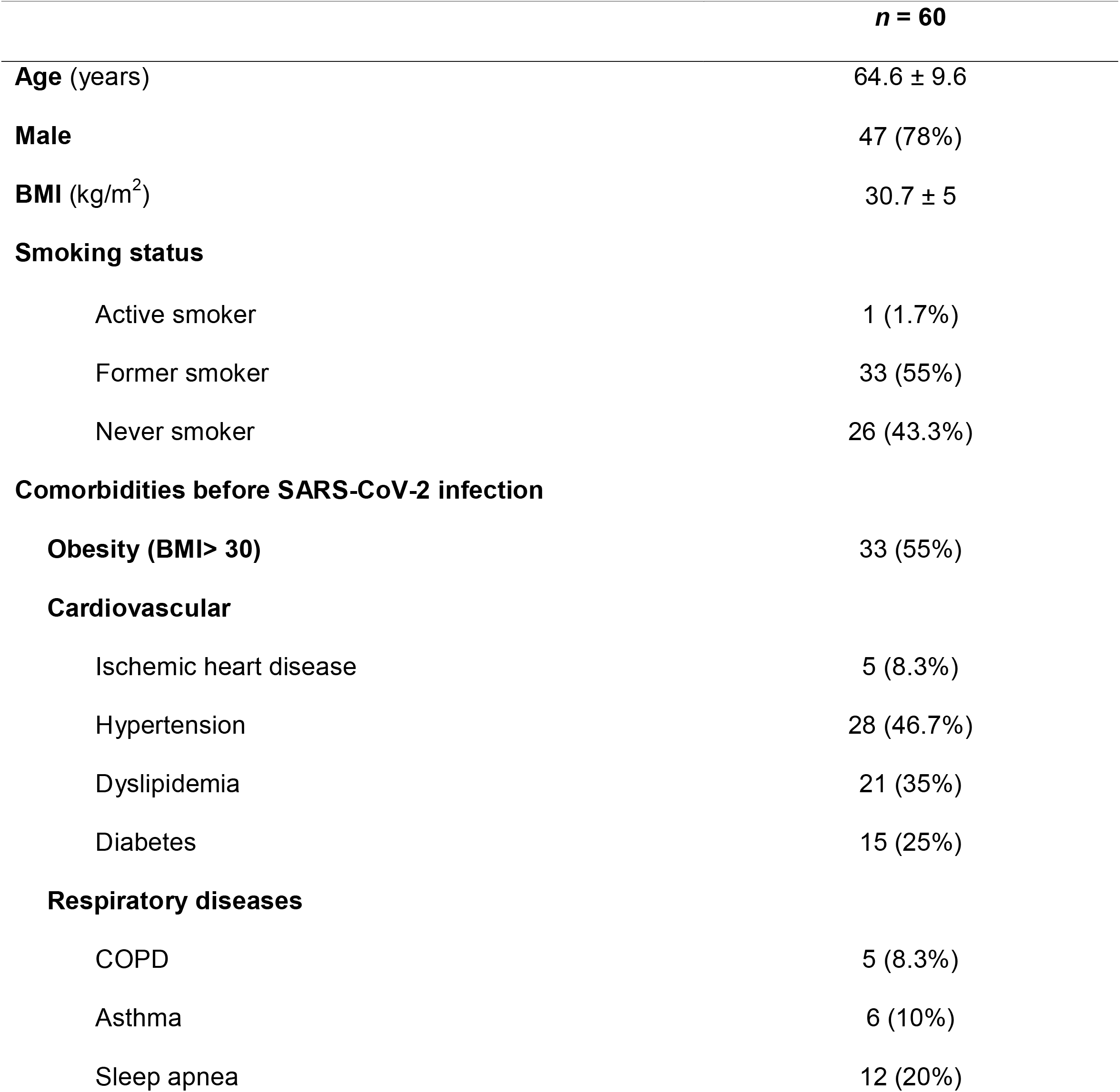
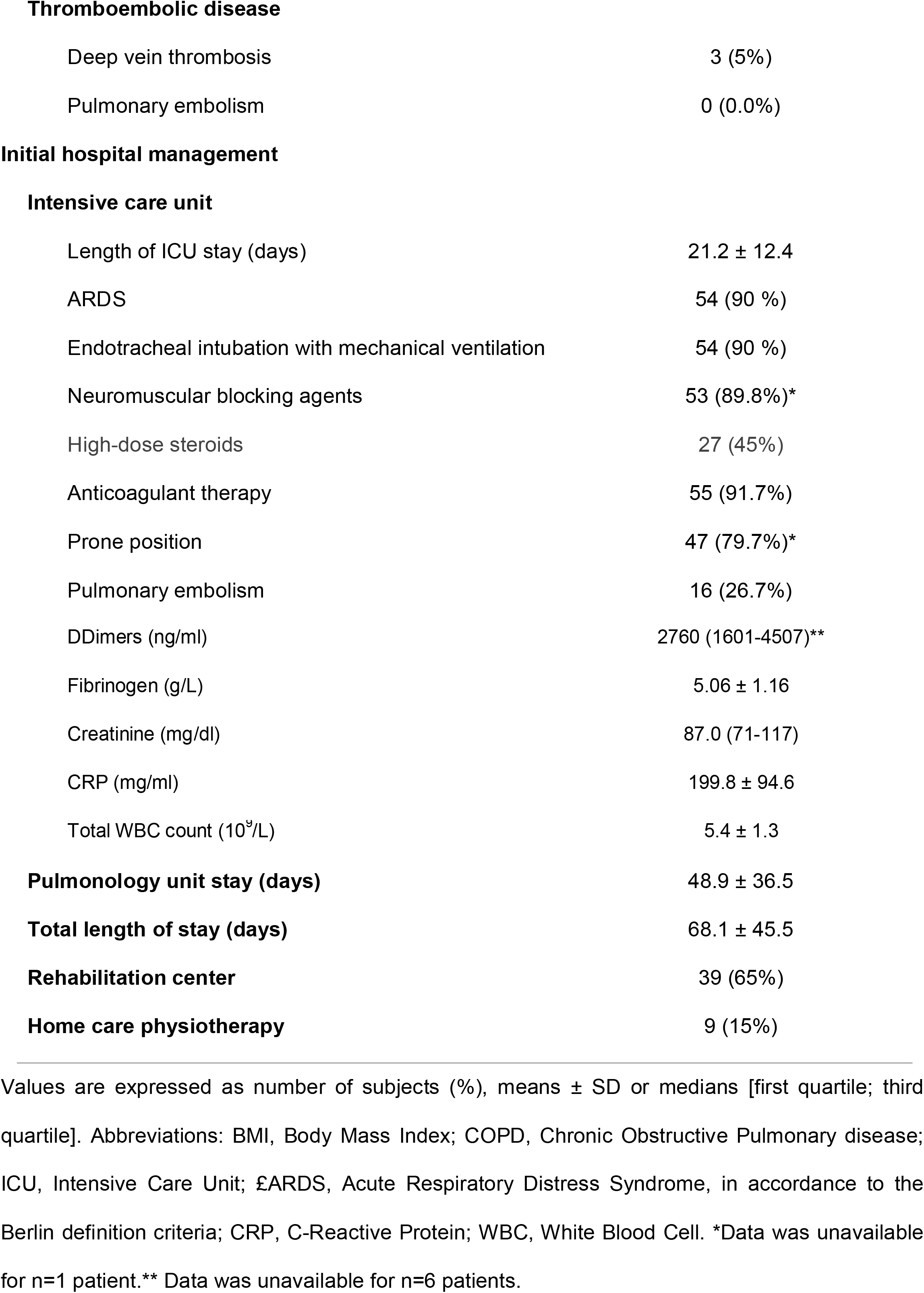
Baseline characteristics of the study population.

### Characteristics of the study population at 12 months follow-up

The clinical, pulmonary function tests and imaging characteristics of the patients at 12 months follow-up are summarized in Table 2. Persistent dyspnea was reported by half of the patients (50%) (mMRC scale ≥ 1). Only a minority of patients had functional pulmonary impairment at rest. Two patients showed a mild restrictive ventilatory pattern, and 4 patients had airflow obstruction, three of them had a previous diagnosis of COPD. Mildly impaired DLCO, defined as Z-score DLCO < −1.64, was present in 6 patients (10%), 4 were already diagnosed with COPD before SARS-CoV2 infection. High resolution computed tomography (HRCT) of the chest showed pulmonary abnormalities in 50 patients (84%). Cardiac evaluation at rest was only proposed to patients that had presented pulmonary embolism during their hospital stay. In these patients, transthoracic echocardiography was within normal limits with no signs of pulmonary hypertension.

**Table 2.**
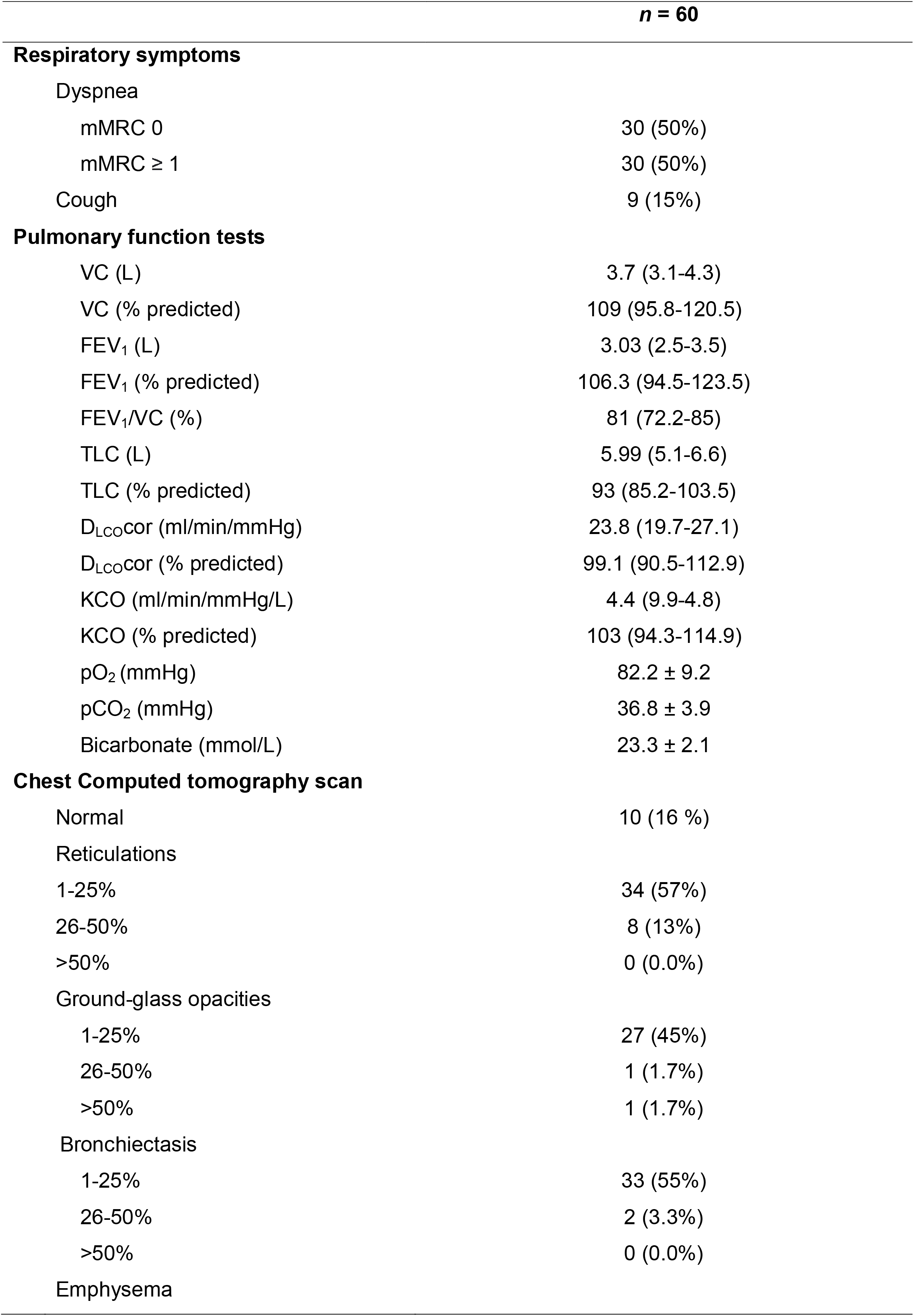
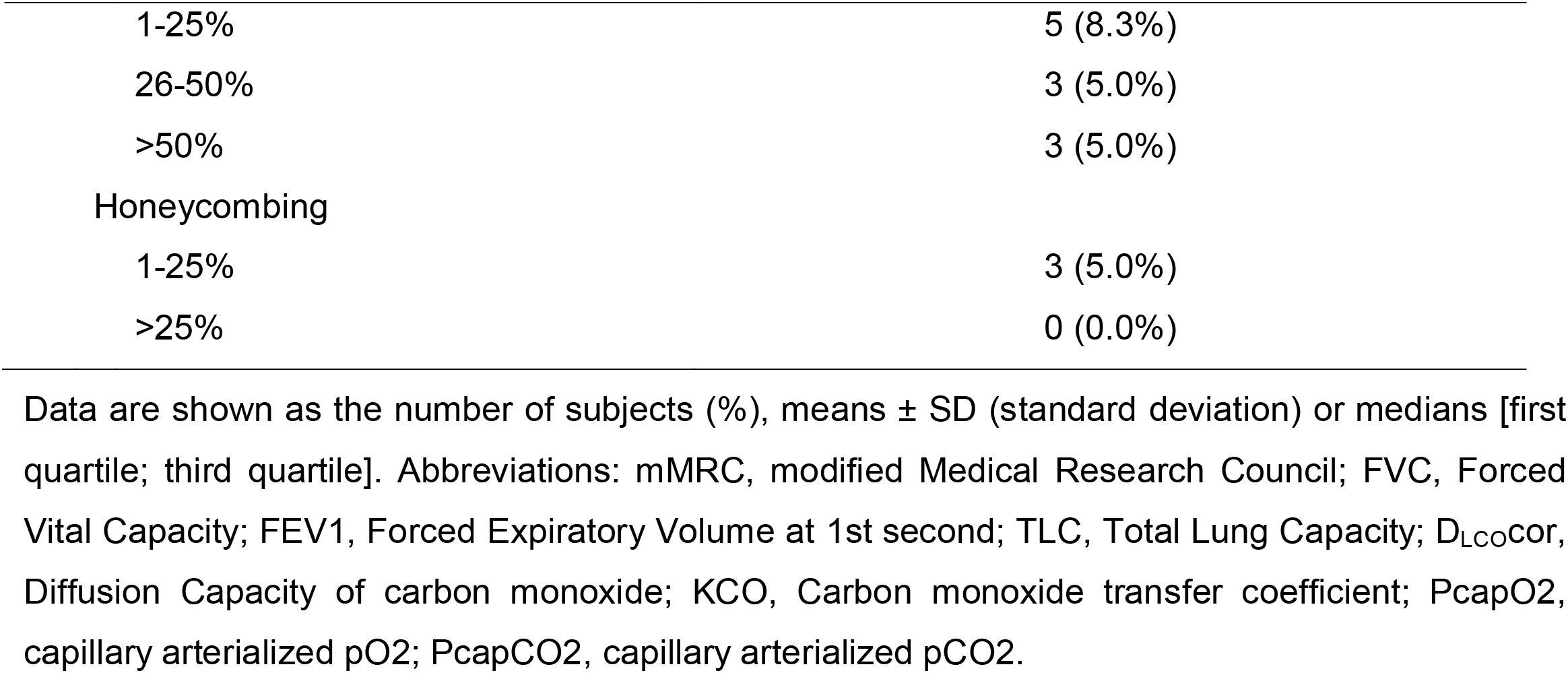
Characteristics of the study population at 12 months follow-up.

|  |  | *n* = 60 |
| --- | --- | --- |
| **Respiratory symptoms** |  |  |
| Dyspnea |  |  |
| mMRC 0 |  | 30 (50%) |
| mMRC ≥ 1 |  | 30 (50%) |
| Cough |  | 9 (15%) |
| **Pulmonary function tests** |  |  |
| VC (L) |  | 3.7 (3.1-4.3) |
| VC (% predicted) |  | 109 (95.8-120.5) |
| FEV<sub>1</sub> (L) |  | 3.03 (2.5-3.5) |
| FEV<sub>1</sub> (% predicted) |  | 106.3 (94.5-123.5) |
| FEV<sub>1</sub>/VC (%) |  | 81 (72.2-85) |
| TLC (L) |  | 5.99 (5.1-6.6) |
| TLC (% predicted) |  | 93 (85.2-103.5) |
| D<sub>LCO</sub>cor (ml/min/mmHg) |  | 23.8 (19.7-27.1) |
| D<sub>LCO</sub>cor (% predicted) |  | 99.1 (90.5-112.9) |
| KCO (ml/min/mmHg/L) |  | 4.4 (9.9-4.8) |
| KCO (% predicted) |  | 103 (94.3-114.9) |
| pO<sub>2</sub> (mmHg) |  | 82.2 ± 9.2 |
| pCO<sub>2</sub> (mmHg) |  | 36.8 ± 3.9 |
| Bicarbonate (mmol/L) |  | 23.3 ± 2.1 |
| **Chest Computed tomography scan** |  |  |
| Normal |  | 10 (16 %) |
| Reticulations |  |  |
| 1-25% |  | 34 (57%) |
| 26-50% |  | 8 (13%) |
| >50% |  | 0 (0.0%) |
| Ground-glass opacities |  |  |
| 1-25% |  | 27 (45%) |
| 26-50% |  | 1 (1.7%) |
| >50% |  | 1 (1.7%) |
| Bronchiectasis |  |  |
| 1-25% |  | 33 (55%) |
| 26-50% |  | 2 (3.3%) |
| >50% |  | 0 (0.0%) |
| Emphysema |  |  |

|  |  |
| --- | --- |
| 1-25% | 5 (8.3%) |
| 26-50% | 3 (5.0%) |
| >50% | 3 (5.0%) |
| Honeycombing |  |
| 1-25% | 3 (5.0%) |
| >25% | 0 (0.0%) |

Data are shown as the number of subjects (%), means ± SD (standard deviation) or medians [first quartile; third quartile]. Abbreviations: mMRC, modified Medical Research Council; FVC, Forced Vital Capacity; FEV1, Forced Expiratory Volume at 1st second; TLC, Total Lung Capacity; $D_{LCOcor}$ , Diffusion Capacity of carbon monoxide; KCO, Carbon monoxide transfer coefficient; PcapO2, capillary arterialized pO2; PcapCO2, capillary arterialized pCO2.

### Cardiopulmonary exercise test (CPET) results at 12 months follow-up

Adequate exercise test efforts were obtained in all the patients analyzed. Table 3 summarizes the main exercise parameters of the study cohort at the anaerobic threshold (AT) and at peak. Most of the patients had an adequate V’O_2_ at AT (median predicted 64.8% [57.2-70.9]). The median peak predicted V’O_2_ was 98 % [87.2-106.3]) (mean peak V’O_2_ 21.7 ± 5.2 mL/min/Kg). Reasons for stopping exercise were leg discomfort in 55% of patients, breathing discomfort in 36.6% patients and both in 5% patients.

**Table 3.**
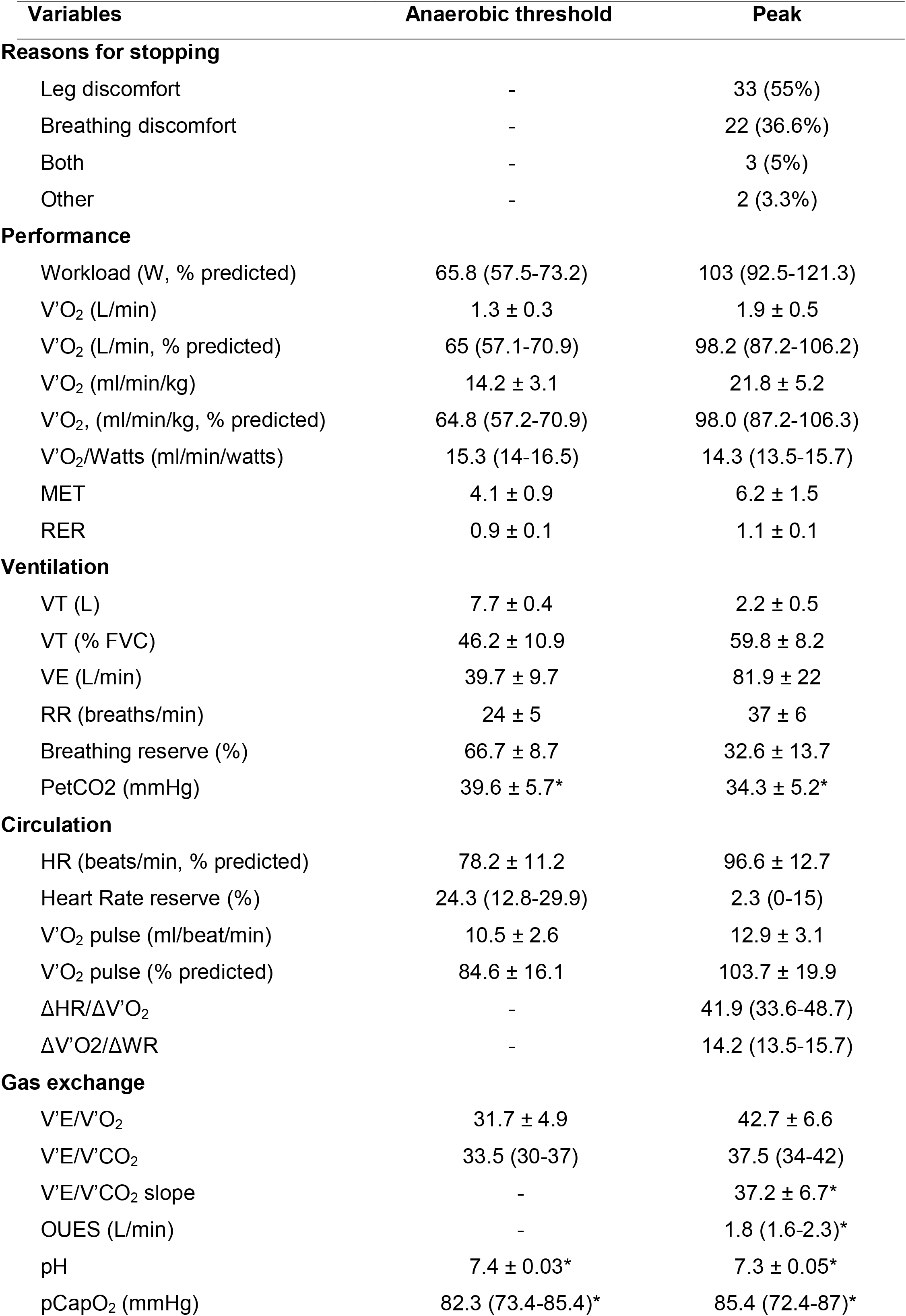
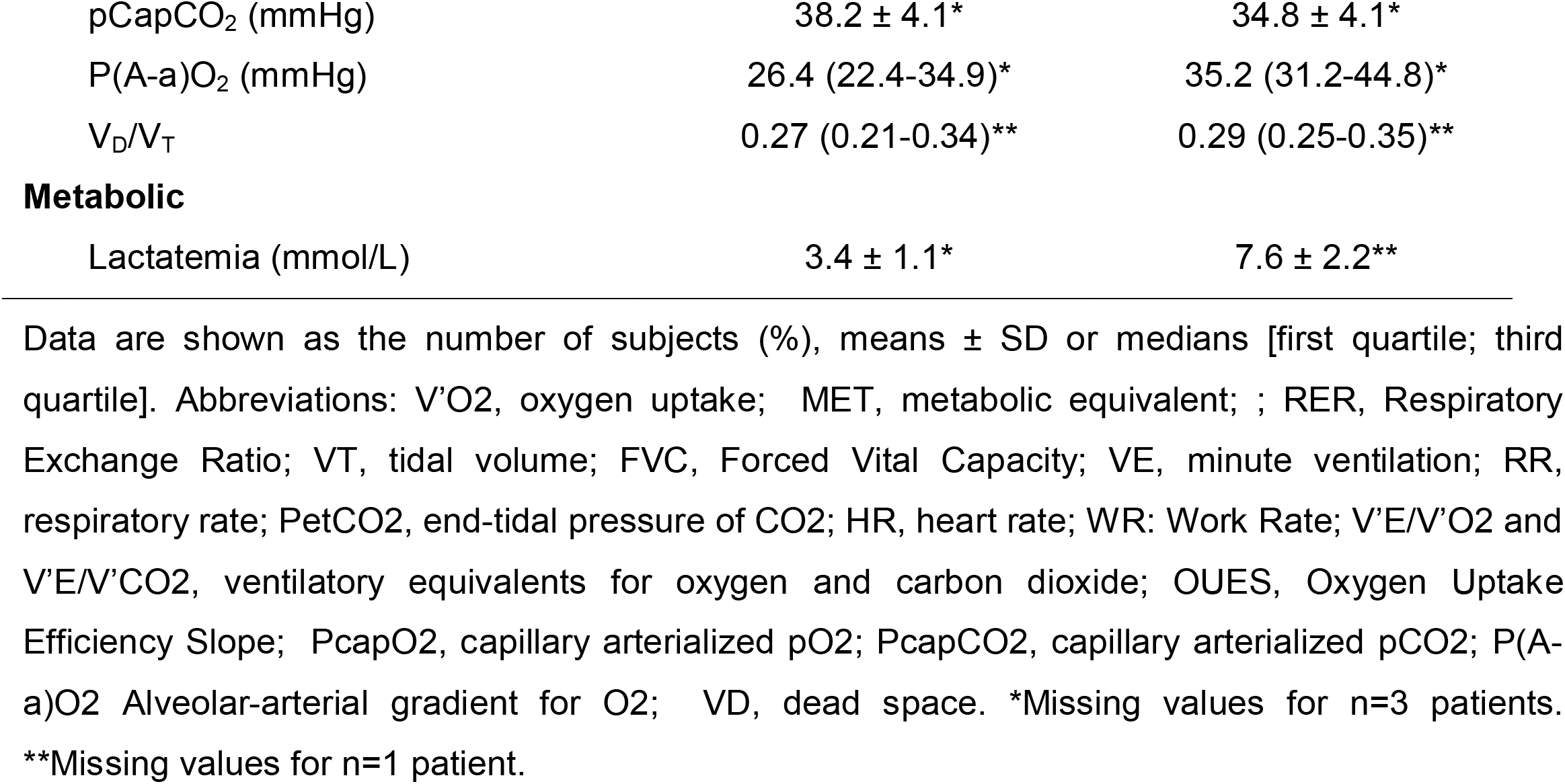
Cardiopulmonary exercise test (CPET) results at 12 months follw-up (n=60).

| Variables | Anaerobic threshold | Peak |
| --- | --- | --- |
| **Reasons for stopping** |  |  |
| Leg discomfort | - | 33 (55%) |
| Breathing discomfort | - | 22 (36.6%) |
| Both | - | 3 (5%) |
| Other | - | 2 (3.3%) |
| **Performance** |  |  |
| Workload (W, % predicted) | 65.8 (57.5-73.2) | 103 (92.5-121.3) |
| V'O<sub>2</sub> (L/min) | 1.3 ± 0.3 | 1.9 ± 0.5 |
| V'O<sub>2</sub> (L/min, % predicted) | 65 (57.1-70.9) | 98.2 (87.2-106.2) |
| V'O<sub>2</sub> (ml/min/kg) | 14.2 ± 3.1 | 21.8 ± 5.2 |
| V'O<sub>2</sub>, (ml/min/kg, % predicted) | 64.8 (57.2-70.9) | 98.0 (87.2-106.3) |
| V'O<sub>2</sub>/Watts (ml/min/watts) | 15.3 (14-16.5) | 14.3 (13.5-15.7) |
| MET | 4.1 ± 0.9 | 6.2 ± 1.5 |
| RER | 0.9 ± 0.1 | 1.1 ± 0.1 |
| **Ventilation** |  |  |
| VT (L) | 7.7 ± 0.4 | 2.2 ± 0.5 |
| VT (% FVC) | 46.2 ± 10.9 | 59.8 ± 8.2 |
| VE (L/min) | 39.7 ± 9.7 | 81.9 ± 22 |
| RR (breaths/min) | 24 ± 5 | 37 ± 6 |
| Breathing reserve (%) | 66.7 ± 8.7 | 32.6 ± 13.7 |
| PetCO<sub>2</sub> (mmHg) | 39.6 ± 5.7* | 34.3 ± 5.2* |
| **Circulation** |  |  |
| HR (beats/min, % predicted) | 78.2 ± 11.2 | 96.6 ± 12.7 |
| Heart Rate reserve (%) | 24.3 (12.8-29.9) | 2.3 (0-15) |
| V'O<sub>2</sub> pulse (ml/beat/min) | 10.5 ± 2.6 | 12.9 ± 3.1 |
| V'O<sub>2</sub> pulse (% predicted) | 84.6 ± 16.1 | 103.7 ± 19.9 |
| ΔHR/ΔV'O<sub>2</sub> | - | 41.9 (33.6-48.7) |
| ΔV'O<sub>2</sub>/ΔWR | - | 14.2 (13.5-15.7) |
| **Gas exchange** |  |  |
| V'E/V'O<sub>2</sub> | 31.7 ± 4.9 | 42.7 ± 6.6 |
| V'E/V'CO<sub>2</sub> | 33.5 (30-37) | 37.5 (34-42) |
| V'E/V'CO<sub>2</sub> slope | - | 37.2 ± 6.7* |
| OUES (L/min) | - | 1.8 (1.6-2.3)* |
| pH | 7.4 ± 0.03* | 7.3 ± 0.05* |
| pCapO<sub>2</sub> (mmHg) | 82.3 (73.4-85.4)* | 85.4 (72.4-87)* |
| pCapCO<sub>2</sub> (mmHg) | 38.2 ± 4.1* | 34.8 ± 4.1* |
| P(A-a)O<sub>2</sub> (mmHg) | 26.4 (22.4-34.9)* | 35.2 (31.2-44.8)* |
| V<sub>D</sub>/V<sub>T</sub> | 0.27 (0.21-0.34)** | 0.29 (0.25-0.35)** |
| Metabolic |  |  |
| Lactatemia (mmol/L) | 3.4 ± 1.1* | 7.6 ± 2.2** |

Data are shown as the number of subjects (%), means ± SD or medians [first quartile; third quartile]. Abbreviations: V'O<sub>2</sub>, oxygen uptake; MET, metabolic equivalent; ; RER, Respiratory Exchange Ratio; VT, tidal volume; FVC, Forced Vital Capacity; VE, minute ventilation; RR, respiratory rate; PetCO<sub>2</sub>, end-tidal pressure of CO<sub>2</sub>; HR, heart rate; WR: Work Rate; V'E/V'O<sub>2</sub> and V'E/V'CO<sub>2</sub>, ventilatory equivalents for oxygen and carbon dioxide; OUES, Oxygen Uptake Efficiency Slope; PcapO<sub>2</sub>, capillary arterialized pO<sub>2</sub>; PcapCO<sub>2</sub>, capillary arterialized pCO<sub>2</sub>; P(A-a)O<sub>2</sub> Alveolar-arterial gradient for O<sub>2</sub>; VD, dead space. \*Missing values for n=3 patients.
\*\*Missing values for n=1 patient.

Circulatory parameters revealed a mean peak predicted oxygen pulse at 103.7 % (± 19.9). Only 8 patients had a mildly decreased O_2_ pulse, 3 of them were under betablockers. The mean peak predicted heart rate was 96.6 % (± 12.7), and the median HR/V’O_2_ slope was in the limit of normal at 41.9 (33.6-48.7).

Mean respiratory equivalents at peak were slightly elevated compared to expected values (V’E/V’O_2_ ratio at 42.7 [± 6.6] and V’E/V’CO_2_ ratio at 37.5 [34-42]) and the mean V’E/V’CO_2_ slope from rest to peak was also slightly skewed to increased values compared to expected values (37.2 ± 6.7) [15]. There was also a trend to a widened median alveolar-arterial O_2_ pressure difference at peak (35.2 mmHg [31.2-44.8]).

#### Predictors of peak oxygen uptake

We next examined the relationship between peak V’O_2_ (ml/min/kg) and variables of interests. Univariate analysis revealed that peak V’O_2_ was strongly correlated to the 6-minute walk test (MWT) distance recorded at 12 months (Figure 1a). As expected, absolute values of lung function test parameters at 12 months (FEV_1_, VC and D_LCO_) were significantly associated to peak V’O_2_ (suppl Table 1).

**Figure 1.**
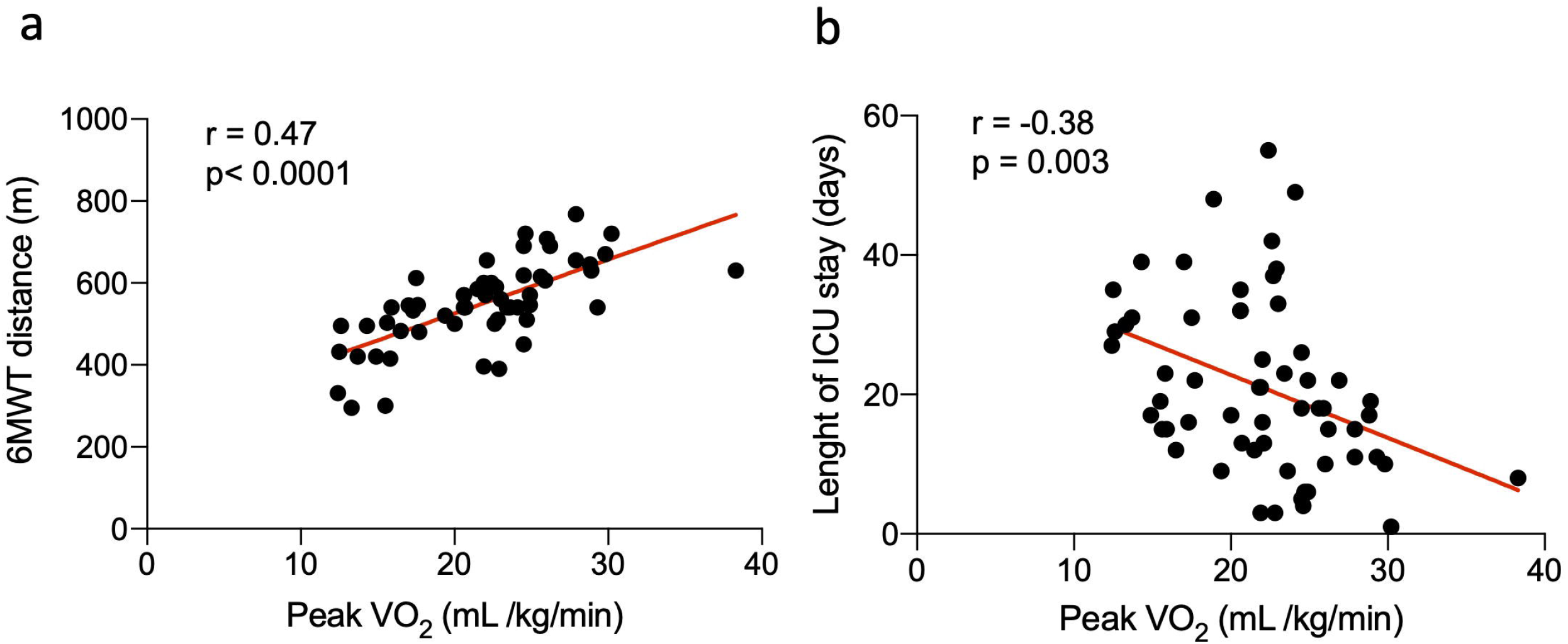
Scatterplot depicting the relationship of peak oxygen uptake (V’O_2_) with a) 6-minute walk test (MWT) distance b) length of ICU stay (days).

Among variables recorded during the management of the acute COVID-19 infection, length of ICU stay showed the most significant correlation with peak V’O_2_ (Figure 1b). Simplified acute physiology score (SAPS II) and length of curarization had a weaker correlation with peak V’O_2_ (suppl Table 1).

In a multiple linear regression analysis, the length of ICU stay remained an independent predictor of V’O_2_ and combined to age, sex, and BMI explained 57% of the variance of V’O_2_ peak at 12 months (Table 4).

**Table 4.**
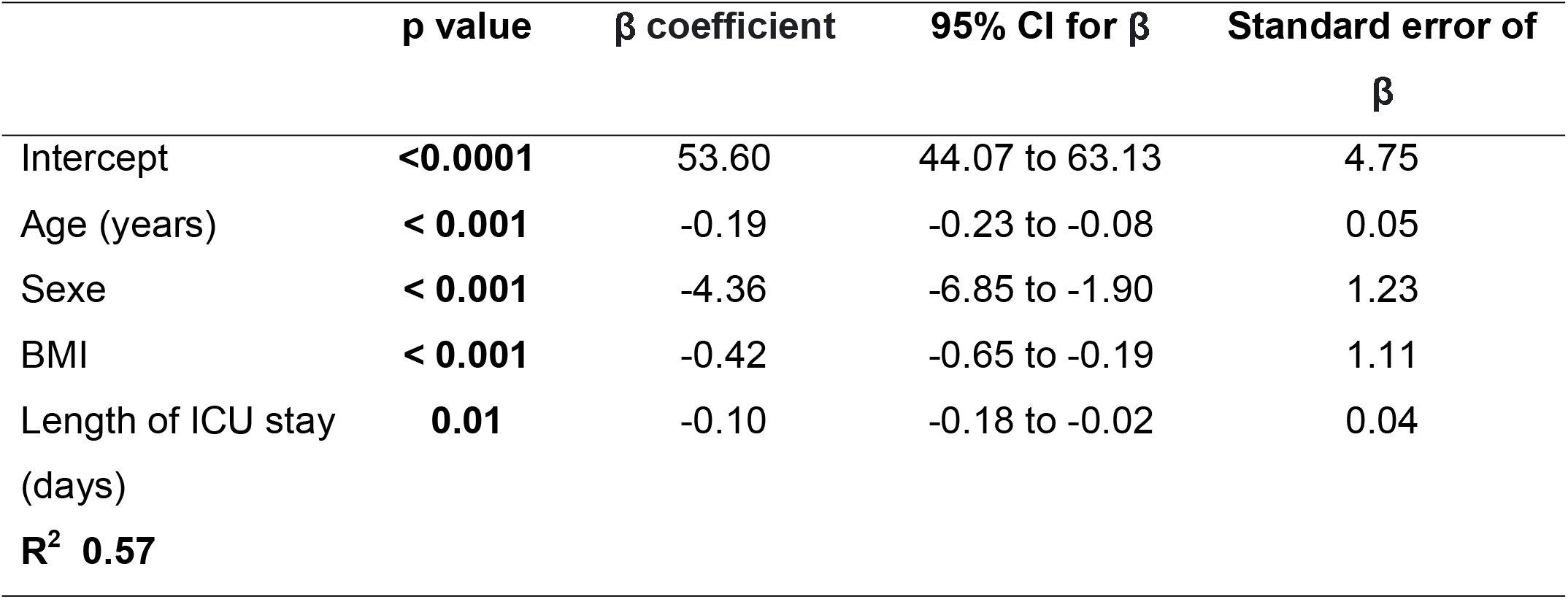
Multiple linear regression identifying factors associated with V’O_2_ peak (ml/kg/min).

#### Comparison of patients with reduced and normal exercise capacity

Twelve patients (20%) had reduced peak exercise capacity (V’O_2_ <85% of predicted) (Table 5). The median peak predicted V’O_2_ (82% [73.9-83.9]) and workload (85.7 % [80.1-96.9]) were only mildly decreased. When compared to patient with preserved peak exercise capacity, patients with reduced capacity had a significantly higher BMI (33.4 ± 6.2 *vs* 30.1 ± 4.5, *p*=0.04) and a significantly longer ICU stay (29.7 ± 13.1 *vs* 19.1 ± 11.3 *p*=0.006). No significant differences were observed between both groups regarding age, other prior comorbidities, pulmonary embolism during ICU stay and respiratory rehabilitation. No significant differences were observed for CT scan abnormalities. However, patients with a reduced exercise capacity had a significantly lower % predicted FEV1 (97.4 ± 16.9 *vs* 110.8 ± 19.7; *p*=0.03), FVC (97.5 ± 17.8 *vs* 110.8 ± 16.9, *p*=0.01) and a lower % predicted D_LCO_ (91.3 (65.6-98.1) *vs* 103 (92.3-114.2); *p*=0.01). Assessment of each individual with limited peak exercise capacity revealed that the primary limitation was ventilatory limitation in 6 patients (50%). Among patients with ventilatory impairment, 5 of them were former smokers and had prior COPD and/or lung emphysema on chest CT. Physical deconditioning was observed for the 6 (50%) others patients.

**Table 5.**
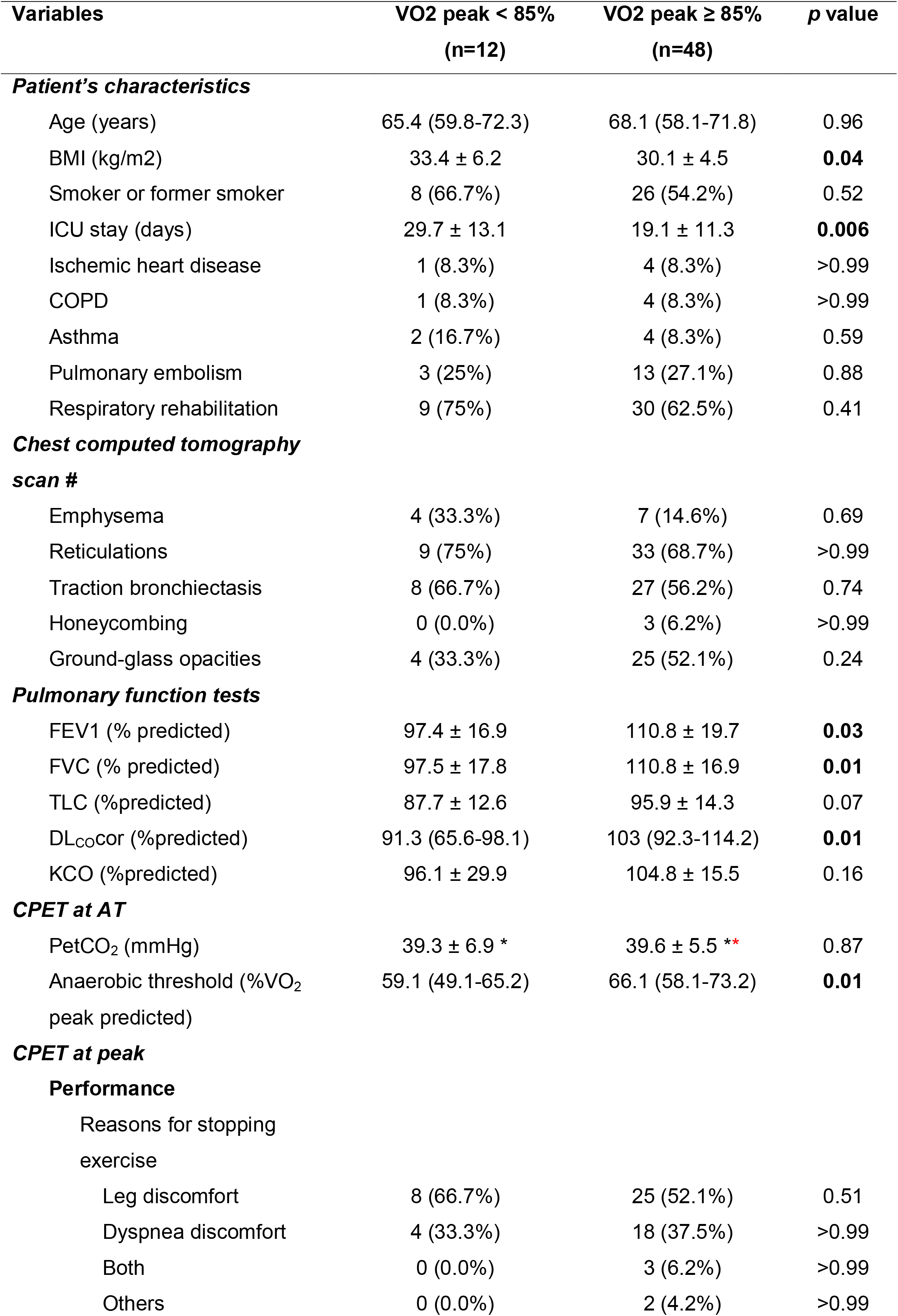
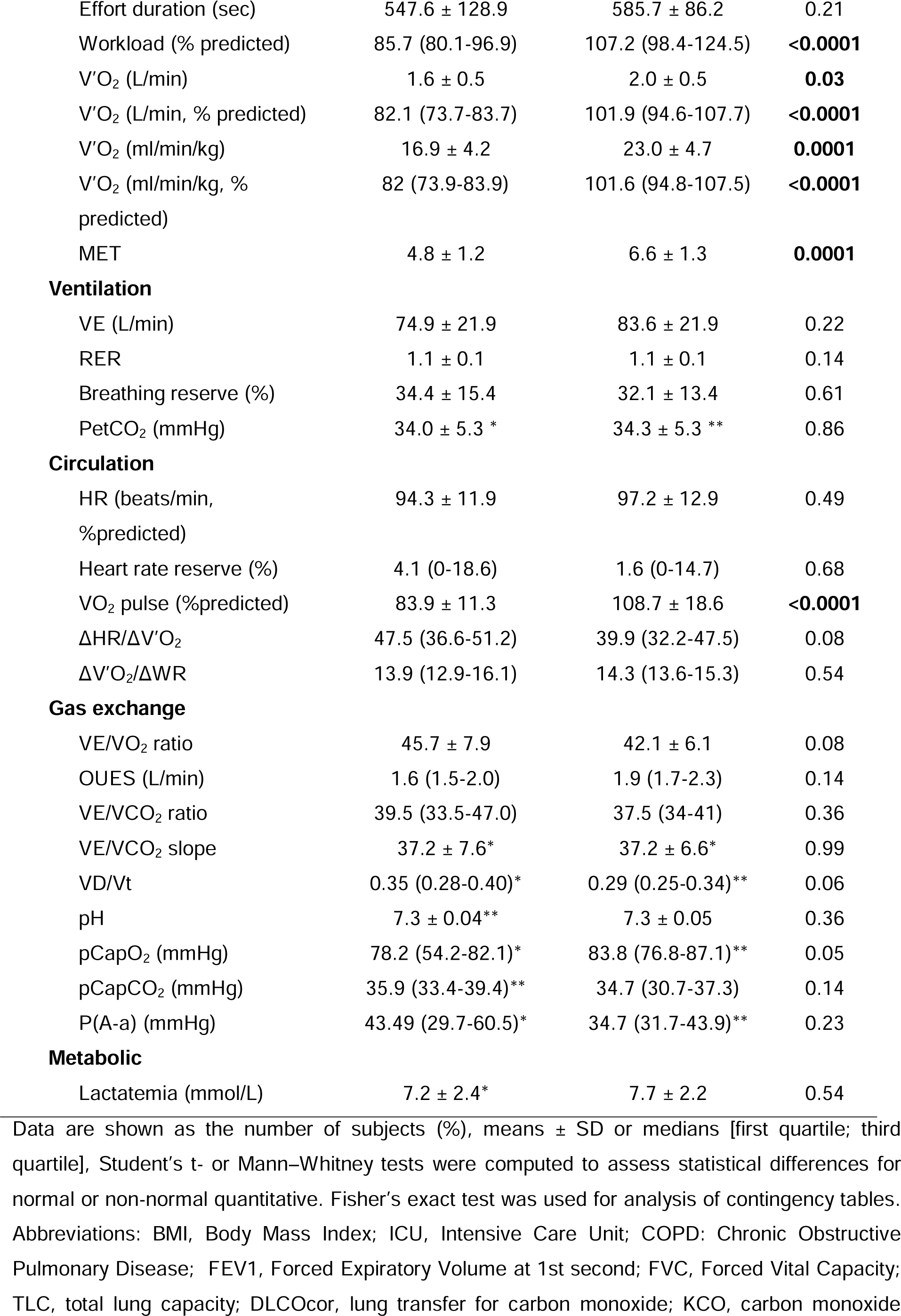
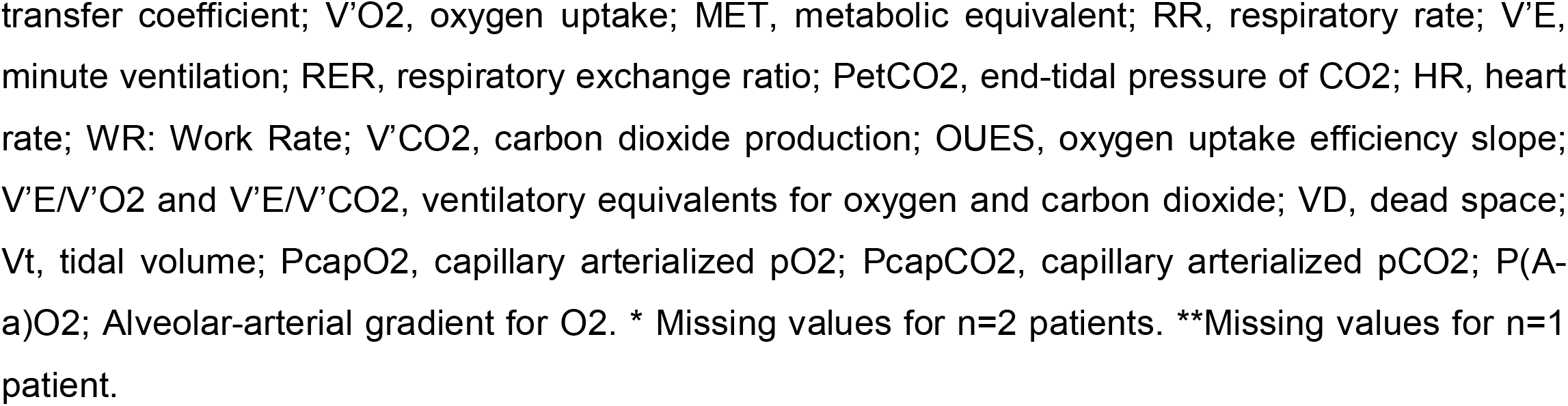
Comparison of patients with reduced and normal exercise capacity.

In the group of patients having an exercise capacity considered within normal limits, the median peak predicted V’O_2_ was 101.6 % [94.8-107.5] (mean of 23.0 ml/kg/min [±4.7]) (Table 5). The main reasons for termination were leg discomfort in 52.1% of patients and dyspnea in 37.5%. Despite having a normal exercise capacity, it was worth noting that patients had increased mean ventilatory equivalents for CO_2_ with a mean peak V’E/V’CO_2_ at 37.5 (34-41) and a mean V’E/V’CO_2_ slope from rest to peak at 37.2 [±6.6]. The median median alveolar-arterial O_2_ pressure difference at peak appeared also widened (34.7 mmHg [31.7-43.9]).

#### Ventilatory efficiency parameters in patients with normal exercise capacity

As we observed that a majority of patients with normal exercise capacity showed elevated mean ventilatory equivalents for CO_2_ during exercise, we next focused on the evolution of ventilatory efficiency parameters in this group.

It was worth noting that at AT, 18 of those 48 patients (37.5%) had a V’E/V’CO_2_ ratio > 35 and 7 patients (14.6%) had even a V’E/V’CO_2_ ratio > 40. At peak exercise, 56.2% (n=27) of patients had a V’E/V’CO_2_ slope > 35 and 41.7% (n=20) had a V’E/V’CO_2_ slope > 40. Anarchical evolution of tidal volume was not observed. In contrast, elevated ventilatory equivalents for CO_2_ were associated with abnormal dead space ventilation. Indeed, an abnormal increase of the physiological dead space from AT to peak was observed in 68.1% (n=32) of the patients with a median VD/Vt of 0.27 (0.21-0.32) at AT and 0.29 (0.25-0.34) at peak (Figure 2a). Moreover, the median peak alveolar-arterial gradient for O_2_ was abnormally elevated (35.2 mmHg [31.2-44.8]) with 48.9 % of patients (n=23) having a P(A-a) ≥ 35 mmHg (Figure 2b). PetCO_2_ at peak was significantly lower in subjects with an abnormal increase of VD/Vt (p=0.001) (Figure 2c).

**Figure 2.**
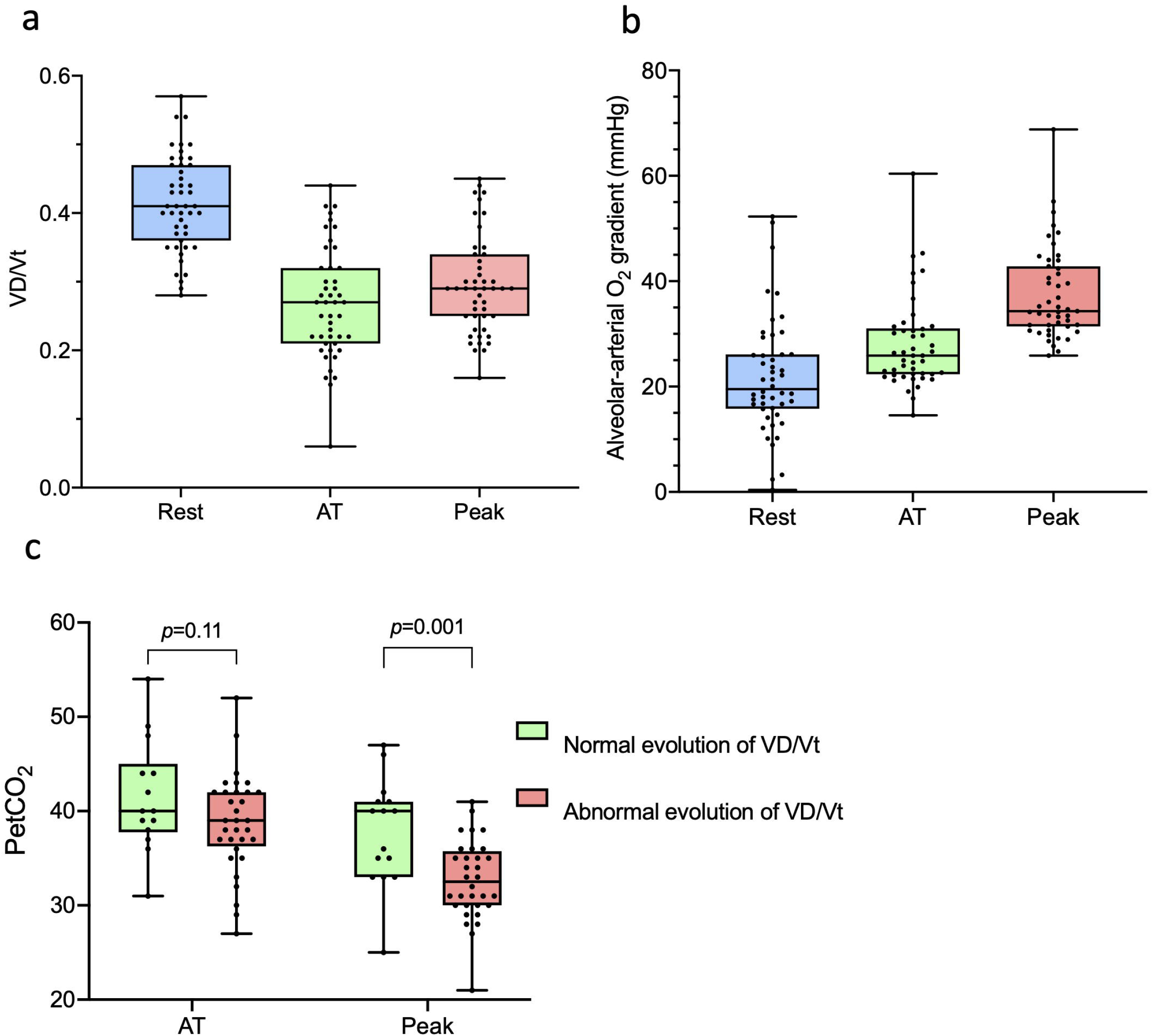
Evolution of ventilatory efficiency parameters from rest to peak in patients with normal exercise capacity (n=48). a) VD/Vt b) alveolar-arterial gradient c) PetCO

Univariate analysis revealed that dead space at peak (VD/Vt) was associated with parameters related to pulmonary exchange capacity at rest (pulmonary diffusing capacity) and during exercise (V’E/V’CO_2_ ratio and slope, PetCO_2_, pO_2_ and alveolar-arterial gradient (Table 6). Notably, dead space at peak was positively correlated to D-Dimer plasma concentration from blood samples collected during ICU stay. Reanalysis after excluding patients with pulmonary embolism during ICU stay did not alter this correlation.

**Table 6.** Results of univariate analysis to identify factors associated with peak VD/Vt in patients with normal exercise capacity (n=48).

| | $r^2$ | p value |
| --- | --- | --- |
| **Patient's characteristics** |  |  |
| Age (years) | 0.13 | **0.01** |
| Sexe | 0.07 | 0.07 |
| BMI (kg/m<sup>2</sup>) | 0.03 | 0.2 |
| Length of ICU stay (days) | 0.24 | 0.10 |
| Pulmonary embolism | 0.01 | 0.3 |
| CRP (mg/ml) | 0.004 | 0.6 |
| Fibrinogen (g/L) | 0.009 | 0.5 |
| D-Dimers (ng/ml) | 0.12 | **0.02** |
| Creatinine (mg/dl) | 0.07 | 0.07 |
| Total WBC count (10<sup>9</sup>/L) | 0.03 | 0.2 |
| **Pulmonary function tests** |  |  |
| FEV1 (% predicted) | 0.08 | 0.05 |
| FEV1 (L) | 0.02 | 0.3 |
| VC (% predicted) | 0.03 | 0.2 |
| VC (L) | 0.002 | 0.7 |
| DL<sub>CO</sub> cor (%predicted) | 0.15 | **0.01** |
| DL<sub>CO</sub> cor (ml/min/mmHg) | 0.06 | 0.09 |
| **CPET at AT** |  |  |
| PetCO<sub>2</sub> | -0.53 | **0.0001** |
| %VO<sub>2</sub> peak predicted | -0.11 | 0.46 |
| **CPET at peak** |  |  |
| V'O<sub>2</sub> peak, ml/kg/min %predicted | -0.17 | 0.25 |
| V'O<sub>2</sub> peak, ml/kg/min |  |  |
| PetCO<sub>2</sub> | -0.43 | **0.003** |
| V'E/VO<sub>2</sub> ratio | 0.48 | **0.0006** |
| V'E/V'CO<sub>2</sub> ratio | 0.58 | **<0.0001** |
| V'E/V'CO<sub>2</sub> slope | 0.53 | **0.0001** |
| V'O<sub>2</sub> pulse (%predicted) | 0.1 | 0.52 |
| pO<sub>2</sub> (mmHg) | 0.13 | **0.01** |
| pCO<sub>2</sub> (mmHg) | 0.004 | 0.8 |
| P(A-a) (mmHg) | 0.46 | **0.01** |

Abbreviations: BMI, Body Mass Index; ICU, Intensive Care Unit; CRP, C-Reactive Protein; WBC, White Blood Cell; FEV1, Forced Expiratory Volume at 1st second; FVC, Forced Vital Capacity; TLC, total lung capacity; DL<sub>CO</sub>cor, lung transfer for carbon monoxide; KCO, carbon monoxide
transfer coefficient; $P_{et}CO_2$ , end-tidal pressure of $CO_2$ ; RER, respiratory exchange ratio; $V'O_2$ , oxygen uptake; $V'E/V'O_2$ and $V'E/V'CO_2$ , ventilatory equivalents for oxygen and carbon dioxide; HR, heart rate; $P_{cap}O_2$ , capillary arterialized $pO_2$ ; $P_{cap}CO_2$ , capillary arterialized $pCO_2$ ; $P(A-a)O_2$ Alveolar-arterial $O_2$ gradient.

Interestingly, patients with persistent dyspnea (n=23) had a significantly higher mean peak dead space (0.32 ±0.07 *vs* 0.28 ±0.06; *p*=0.04) and a higher widening of their mean peak alveolar-arterial gradient (40.9 ± 9.8 vs 34.2 ± 5.9; *p*=0.006) (Suppl Table 2). There was no significant difference for the mean peak VE/VCO_2_ ratio and the mean VE/VCO_2_ slope. Performance and circulatory parameters during exercise were also similar between both groups. As expected, breathlessness was the predominant symptom that resulted in test termination in patients with persistent dyspnea. Patients with persistent dyspnea were slightly older. No differences were observed for any other parameters recorded during initial hospitalization including the presence of pulmonary embolism between patients with and without persistent dyspnea. Lung function at rest was also similar at 12 months between both groups (Suppl Table 2).

## Discussion

This prospective study assessed cardiopulmonary exercise performance in 60 patients, 12 months after a critical COVID-19 infection during the first wave that required initial ICU management. The patients of our cohort presented well-established risk factors for severe COVID-19 such as advanced age, a predominance of male sex and a high BMI [16]. As expected, hypertension and dyslipidemia were the most frequent chronic comorbidities.

Despite the severity of the initial clinical presentation, exercise capacity assessed by CPET were within normal limits in most of the patients 12 months after the acute infection. Impairment was predominantly related to persistent deconditioning or prior respiratory co-morbidities. These results confirm previous studies assessing exercise capacity by CPET 3 to 6 months after hospital release and reporting that remaining exercise limitation after COVID-19 is primarily related to physical deconditioning rather than to physiological impairment [5, 6, 17, 18]. Thus, recovery of physical capacities after a critical COVID-19 infection appears better than in patients with other ARDS etiologies [19, 20].

In our study, 12 months after the acute infection, the length of ICU stay was still an independent predictor of peak V’O_2,_ including the patients that had recovered a peak V’O_2_ considered within normal limits. Even if other studies have already reported associations between length of ICU stay after COVID-19 and peak V’O_2_ [21, 22], we were surprised that this association remained still true several months after hospital release. This was even more unexpected as all our patients had received early physiotherapy management in the acute hospital setting followed by either inpatient rehabilitation or extensive physiotherapy for several weeks at home for most of them.

Another intriguing observation in our study was that many patients having a peak V’O_2_ within normal limits and normal rest lung function, showed ventilatory inefficiency during exercise, with increased V’E/V’CO_2_ ratios at AT and at peak associated with an increased V’E/V’CO_2_ slope. Ventilatory inefficiency after acute COVID-19 has already been reported but mainly attributed to dysfunctional breathing with inappropriate hyperventilation suggestive of post-traumatic syndrome [23]. In contrast to our study, these studies enrolled predominantly patients having had mild COVID-19 [21, 24-26].

In our patients, there was no evidence of exaggerated hyperventilatory response. Indeed, we did not see abnormal respiratory alkalosis, nor anarchical evolution of tidal volume. However, ventilatory inefficiency was associated in our study with increased physiological dead space ventilation. Indeed, we observed that nearly two-thirds of the patients with a normal peak predicted V’O_2_ exhibited an increase of the VD/Vt ratio between AT and peak exercise. Ventilatory inefficiency without hyperventilation syndrome has been suggested by other groups but these studies included patients with a range of disease severity combining data from outpatients, hospitalized patients, and those who had required admission to the ICU [22, 27]. Of interest, Ambrosino *et al*. identified in a study that included mostly severe-to-critical COVID-19 patients without any prior history of cardiovascular or pulmonary disease shortly after hospital release, higher VE/VCO_2_ ratios and VE/VCO_2_ slopes and a lower VD/Vt decrease among patients with reduced exercise capacity [28].

In healthy individuals, VD/Vt decreases usually during exercise as Vt increases several folds and to a much greater extent than the small increase in VD [29]. An increase of the physiological dead space ventilation during exercise is usually considered as abnormal and may suggest the presence of several cardiac and pulmonary disorders. However, increasing VD/VT during exercise is mostly sensitive for pulmonary vascular disease [30-32]. As in our study the majority of patients with normal exercise capacity had normal rest pulmonary function and no cardiovascular abnormalities, the increase of physiological dead space ventilation associated to a low peak PetCO_2_ may point to pulmonary vascular disease.

It is now apparent that SARS-CoV-2 infection induces endothelial cell dysfunction with systemic inflammatory response resulting in a prothrombotic state manifesting especially with microthrombosis [1]. Notably, in our study, peak VD/Vt values at 12 months were positively correlated to peak D-Dimers plasma concentrations from blood samples collected during ICU stay. D-Dimers are a strong biomarker for hypercoagulability and thrombotic events and can be linked to endothelial dysfunction reported during acute COVID-19 [33, 34]. Therefore, the observed ventilatory inefficiency in our patients may point to infra-clinical pulmonary vasculopathy sequelae due to lung micro-thrombotic events during acute SARS-CoV-2 infection. In a study aiming to quantify endothelial alterations in 23 patients with moderate to critical COVID-19, sublingual video microscopy confirmed microcirculatory alterations that were closely associated with D-Dimer levels [35]. More recently, in a cohort of severe-to-critical COVID-19 patients, Ambrosino *et al*. showed that persistent endothelial dysfunction explored by ultrasound assessment of endothelium-dependent flow-mediated dilation (FMD) was correlated to ventilatory inefficiency parameters during CPET in a subgroup of patients [28].

Half of our patients, including those who had an exercise capacity within normal limits, still complained of persistent dyspnea. A recent study evaluating the health-related quality of life and persistent symptoms in critically ill COVID-19 patients at twelve months identified a similar proportion with 58.4% of patients with persistent mild dyspnea that was weakly correlated with both DLCO and length of invasive mechanical ventilation [36]. In our study, we found no clear association between dyspnea, length of ICU stay, effort capacity or rest lung function parameters. However, patients with persistent dyspnea had significantly higher mean peak dead space associated to a higher widening of their mean peak alveolar-arterial gradient during exercise.

Some potential limitations of our study should be noted. Our study was conducted in a single center. There was also a missing baseline assessment of cardiopulmonary function at rest before SARS-CoV-2 infection in our patients. Even if nearly all the patients with normal exercise capacity and ventilatory inefficiency at exercise had spirometry and predicted DLCO within normal limits, it would have been interesting to compare the results with matched controls. Indeed, more than half of our study cohort were smokers or former smokers. Moreover, we were not able to measure the diffusing capacities of the lung for nitric oxide (NO) which combined to D_LCO_ would have been useful to evaluate more precisely the pulmonary vascular implication. Finally, all patients of our cohort were treated according to local standards at the time of the first wave of COVID-19. Consequently, only 45% of them received corticosteroids but notably all of the patients were precociously anticoagulated during their ICU stay.

## Conclusion

In the current study we report that 12 months after critical COVID-19 most of the patients had a peak V’O_2_ considered within normal limits. The length of the ICU stay remained a significant predictor of peak V’O_2_ and therefore prolonged cardiopulmonary rehabilitation and exercise activity should be encouraged in this population. Notably, more than half of the patients who had a normal exercise capacity were still complaining of persistent dyspnea and two-thirds showed abnormal ventilatory efficiency during exercise suggestive of pulmonary vasculopathy. These findings may raise the question of a prolonged antithrombotic therapy in the management of critical post-COVID-19. Further studies, based on invasive hemodynamic measurements during exercise are required to clarify our observations.

## Data Availability

All data produced in the present study are available upon reasonable request to the authors

## Figure Legends

**Suppl Figure**. Flowchart of the prospective cohort study.

