## Supplementary figures and images for "Evaluation of long-term sequelae by cardiopulmonary exercise testing 12 months after hospitalization for critical COVID-19"

### Supplemental Figure 1

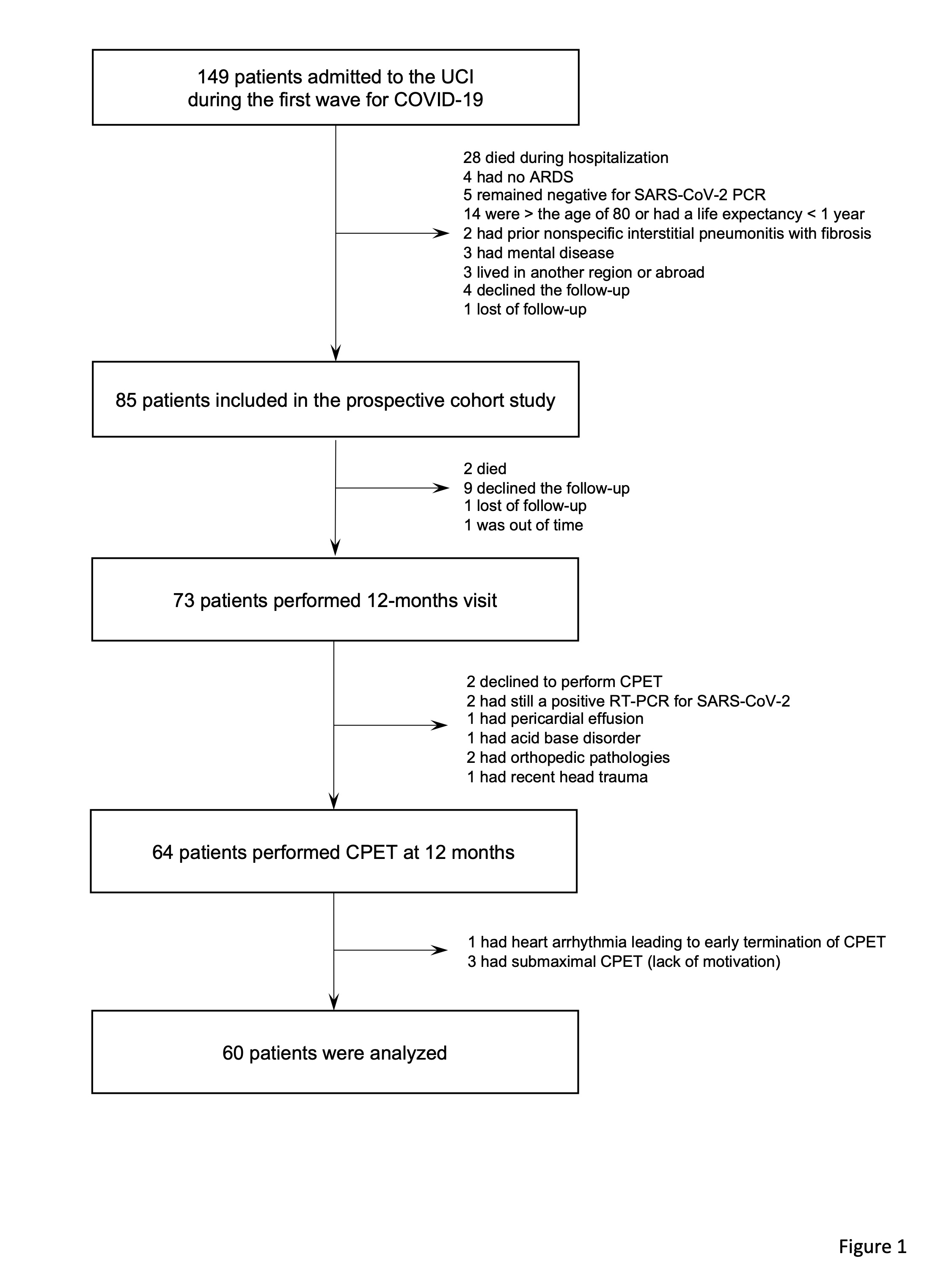
