## Supplemental Table 1 for "Evaluation of long-term sequelae by cardiopulmonary exercise testing 12 months after hospitalization for critical COVID-19"

**Suppl Table 1.** Results of univariate analysis to identify factors associated with V’O_2_ peak (ml/kg/min).

|  | **r^2^** | **p value** |
| --- | --- | --- |
| Age (years) | 0.14 | **0.003** |
| Sexe | 0.20 | **0.0003** |
| Weight (kg) | 0.05 | 0.07 |
| BMI | 0.25 | **< 0.001** |
| Tobacco status | 0.005 | 0.4 |
| SAPS II | 0.02 | **0.03** |
| SOFA score | 0.07 | 0.2 |
| Total length of Curarization (days) | 0.10 | **0.02** |
| High-dose steroids | 0.03 | 0.1 |
| Length of ICU stay (days) | 0.14 | **0.003** |
| Total length of stay (days) | 0.07 | **0.04** |
| Pulmonology unit length of stay (days) | 0.04 | 0.09 |
| Pulmonary embolism | 0.004 | 0.6 |
| CRP (ml/L) | 0.0005 | 0.8 |
| Total WBC count (10^9^/L) | 0.05 | 0.07 |
| Ferritine (ng/ml) | 0.0006 | 0.8 |
| DDimers (ng/ml) | 0.03 | 0.1 |
| Fibrinogen (g/L) | 0.05 | 0.07 |
| Platelet count (109/L) | 0.01 | 0.3 |
| Creatinine (mg/dl) | 0.004 | 0.6 |
| VC (L) | 0.24 | **< 0.0001** |
| VC (% predicted) | 0.01 | 0.3 |
| FEV1 (L) | 0.26 | **< 0.0001** |
| FEV1 (% predicted) | 0.007 | 0.8 |
| TLC (L) | 0.19 | **0.004** |
| TLC (% predicted) | 0.001 | 0.8 |
| DLCO (ml/min/mmHg) | 0.22 | **0.0001** |
| DLCO (% predicted) | 0.01 | 0.3 |
| KCO (ml/min/mmHg/L) | 0.008 | 0.5 |
| KCO (% predicted) | 0.004 | 0.6 |
| Six-minute walk test | 0.46 | **< 0.0001** |
| Rest Heart Rate | 0.005 | 0.5 |
| Rest Saturation (%) | 0.0001 | 0.9 |
| PaO_2_ at rest (mmHg) | 0.001 | 0.6 |
| PaCO_2_ at rest (mmHg) | 0.004 | 0.8 |

Abbreviations: BMI, body mass index; FVC, Forced Vital Capacity; FEV_1_, Forced Expiratory Volume at 1st second; TLC, total lung capacity; D_LCO_, Diffusion Capacity of carbon monoxide; KCO, carbon monoxide transfer coefficient, PcapCO_2_, capillary arterialized pCO_2_; PcapO_2_, capillary arterialized pO_2_. SAPS II, simplified acute physiology score. SOFA score, sequential organ failure assessment score.
