## Supplemental Table 2 for "Evaluation of long-term sequelae by cardiopulmonary exercise testing 12 months after hospitalization for critical COVID-19"

**Suppl Table 1.** Comparison of patients with and without persistent dyspnea and normal exercise capacity

| **Variables** | | | | **mMRC = 0**  **(n = 25)** | **mMRC ≥ 1**  **(n = 23)** | **p-value** |
| --- | --- | --- | --- | --- | --- | --- |
| ***Patient’s characteristics*** | | | |  |  |  |
|  | Age (years) | | | 64.4 (55.9-69.1) | 69.5 (66.8-72.9) | **0.02** |
|  | BMI (kg/m2) | | | 29.8 ± 3.6 | 30.3 ± 5.4 | 0.70 |
|  | Smoker or former smoker | | | 11 (44%) | 15 (65.2%) | 0.16 |
|  | ICU stay (days) | | | 17.2 ± 9.4 | 21.0 ± 13.0 | 0.24 |
|  | Pulmonary embolism | | | 6 (24%) | 5 (21.7%) | >0.99 |
|  | Respiratory rehabilitation | | | 15 (60%) | 15 (65.2%) | 0.77 |
| ***Pulmonary function tests*** | | | |  |  |  |
|  | FEV1 (% predicted) | | | 115.6 ± 18.7 | 105.5 ± 19.9 | 0.07 |
|  | FVC (% predicted) | | | 114.6 ± 17.2 | 106.5 ± 16.01 | 0.09 |
|  | TLC (%predicted) | | | 95 (89-101) | 98 (88-104) | 0.87 |
|  | D_LCO_cor (%predicted) | | | 103.9 ± 14.8 | 103.2 ± 20.7 | 0.89 |
|  | KCO (%predicted) | | | 105.1 ± 10.3 | 104.5 ± 20.04 | 0.88 |
| ***CPET at AT*** | | | |  |  |  |
|  | Anaerobic threshold (%VO_2_ peak predicted) | | | 66.0 (55.4-75.1) | 64.0 (55.3-67.6) | 0.44 |
|  | PetCO_2_ (mmHg) | | | 40.6 ± 5.7 | 38.6 ± 5.1* | 0.23 |
| ***CPET at peak*** | | | |  |  |  |
|  | **Performance** | | |  |  |  |
|  | | Dyspnea, Borg | | 8 (5.5-10.0) | 8 (7-10)* | 0.48 |
|  | | Reasons for stopping exercise | |  |  |  |
|  | | | Leg discomfort | 18 (72%) | 7 (30.4%) | **0.008** |
|  | | | Dyspnea discomfort | 5 (20.0 %) | 13 (56.5%) | **0.01** |
|  | | | Both | 1 (4.0%) | 2 (8.7%) | 0.60 |
|  | | | Others | 1 (4.0%) | 1 (4.3%) | >0.99 |
|  | | Effort duration (seconds) | | 574.4 ± 103.7 | 598.2 ± 59.3 | 0.33 |
|  | | Workload (% predicted) | | 114.2 ± 25.2 | 112.3 ± 21.1 | 0.78 |
|  | | V’O_2_ peak, ml/kg/min %predicted | | 102.7 (96.3-111.8) | 100.9 (93.2-106.9) | 0.37 |
|  | **Ventilation** | | |  |  |  |
|  | | VE (L/min) | | 85.4 ± 20.7 | 81.6 ± 23.5 | 0.56 |
|  | | RER | | 1.1 ± 0.1 | 1.11 ± 0.1 | 0.69 |
|  | | Breathing reserve (%) | | 34.7 ± 12.7 | 29.3 ± 14.0 | 0.16 |
|  | | PetCO_2_ (mmHg) | | 34.9 ± 5.80 | 33.8 ± 4.8* | 0.48 |
|  | **Circulation** | | |  |  |  |
|  | | HR (beats/min, %predicted) | | 97.7 ± 13.3 | 96.7 ± 12.9 | 0.79 |
|  | | Heart rate reserve (%) | | 0.6 (0.0-14.3) | 2 (0.0-15.2) | 0.82 |
|  | | VO_2_ pulse (%predicted) | | 104.7 ± 24.7 | 107.2 ± 19.8 | 0.69 |
|  | | ΔHR/ΔV’O_2_ | | 43.8 (31.2-49.2) | 40.3 (33.3-47.6) | 0.83 |
|  | | ΔV’O_2_/ΔWR | | 14.14 (13.5-15.2) | 14.70 (13.6-15.8) | 0.52 |
|  | **Gas exchange** | | |  |  |  |
|  | | VE/VO_2_ ratio | | 41.4 ± 6.9 | 42.6 ± 5.1 | 0.49 |
|  | | OUES | | 1.9 (1.6-2.5) | 1.9 (1.7-2.3)* | 0.69 |
|  | | VE/VCO_2_ ratio | | 37.3 ± 6.4 | 38.7 ± 5.1 | 0.38 |
|  | | VE/VCO_2_ slope | | 37.4 ± 7.1 | 37.1 ± 6.1* | 0.89 |
|  | | VD/Vt | | 0.28 (±0.06) | 0.32 (±0.07)* | **0.04** |
|  | | pH | | 7.3 ± 0.05 | 7.3 ± 0.04 | 0.44 |
|  | | pCapO_2_ (mmHg) | | 84.6 ± 7.5* | 77.2 ± 1.5 | **0.01** |
|  | | pCapCO_2_ (mmHg) | | 34.1 ± 3.9 | 34.8 ± 4.3 | 0.51 |
|  | | P(A-a) (mmHg) | | 34.2 ± 5.9* | 40.9 ± 9.8 | **0.006** |
|  | **Metabolic** | | |  |  |  |
|  | | Lactatemia (mmol/L) | | 8.4 (6.8-9.5) | 7.4 (5.7-8.8) | 0.25 |

Data are shown as the number of subjects (%), means ± SD or medians [first quartile; third quartile], Student’s t- or Mann–Whitney tests were computed to assess statistical differences for normal or non-normal quantitative. Fisher’s exact test was used for analysis of contingency tables. Abbreviations: BMI, Body Mass Index; ICU, Intensive Care Unit;  FEV1, Forced Expiratory Volume at 1st second; FVC, Forced Vital Capacity; TLC, total lung capacity; DL_CO_cor, lung transfer for carbon monoxide; KCO, carbon monoxide transfer coefficient; V’O_2_, oxygen uptake; VE, minute ventilation; RER, respiratory exchange ratio; PetCO_2_, end-tidal pressure of CO_2_; HR, heart rate; WR: Work Rate; OUES, oxygen uptake efficiency slope; V’CO_2_, carbon dioxide production; V’E/V’O_2_ and V’E/V’CO_2_, ventilatory equivalents for oxygen and carbon dioxide; VD: Dead Volume; VT, tidal volume; PcapO_2_, capillary arterialized pO_2_; PcapCO_2_, capillary arterialized pCO_2_; P(A-a)O_2_ Alveolar-arterial gradient for O_2_. *Missing values for n=1 patient.
